# One vaccine to counter many diseases? Modelling the economics of oral polio vaccine against child mortality and COVID-19

**DOI:** 10.1101/2022.01.19.22269560

**Authors:** Angela Y. Chang, Peter Aaby, Michael S. Avidan, Christine S. Benn, Stefano M. Bertozzi, Lawrence Blatt, Konstantin Chumakov, Shabaana A. Khader, Shyam Kottilil, Madhav Nekkar, Mihai G. Netea, Annie Sparrow, Dean T. Jamison

## Abstract

**Background:** Recent reviews summarize evidence that some vaccines have heterologous or non-specific effects (NSE), potentially offering protection against multiple pathogens. Numerous economic evaluations examine vaccines’ pathogen-specific effects, but we have found only two economic evaluations of NSE. This paper starts to fill this gap by reporting economic evaluations of the NSE of oral polio vaccine (OPV) against under-five mortality and COVID-19.

**Methods:** We studied two settings: (1) reducing child mortality in a high-mortality setting (Guinea-Bissau) and (2) preventing COVID-19 in India. In the former, the intervention involves three annual campaigns in which children receive OPV incremental to routine immunization. In the latter, a susceptible-exposed-infectious-recovered model was developed to estimate the population benefits of two scenarios, in which OPV would be co-administered alongside COVID-19 vaccines. Incremental cost-effectiveness and benefit-cost ratios were modelled for ranges of intervention effectiveness estimates to supplement the headline numbers and account for heterogeneity and uncertainty.

**Results:** For child mortality, headline cost-effectiveness was $650 per child death averted. For COVID-19, assuming OPV had 20% effectiveness, incremental cost per death averted was $23,000-65,000 if it were administered simultaneously with a COVID-19 vaccine less than 200 days into a wave of the epidemic. If the COVID-19 vaccine availability were delayed, the cost per averted death would decrease to $2600-6100. Estimated benefit-to-cost ratios vary but are consistently high.

**Conclusion:** Economic evaluation suggests the potential of OPV to efficiently reduce child mortality in high mortality environments. Likewise, within a broad range of assumed effect sizes OPV could play an economically attractive role against COVID-19.

## Introduction

Vaccination to induce acquired, highly effective, pathogen-specific immunity now covers most of the world. Numerous studies have examined the economic impact of existing vaccines against the pathogens they target.(1,2) Less known, but well demonstrated scientifically, is the fact that live attenuated vaccines (LAVs) can also induce broader innate immune protection against unrelated pathogens.(3) These non-specific heterologous effects of LAVs – such as the Bacillus Calmette-Guérin (BCG) vaccine against tuberculosis, measles-containing vaccines, and oral polio vaccines (OPV) – have led to reductions in mortality and morbidity by more than can be explained from prevention of the targeted disease alone.(4) Only two studies have undertaken economic analyses that explicitly incorporates the heterologous effects of pathogen-specific vaccines. Byberg et al. concluded that the heterologous effects were more important in determining cost-effectiveness than the measles-specific effect, while Thompson et al. assumed there is insufficient evidence for a non-specific heterologous effect of OPV against COVID-19 in the United States.(5,6) We aim to address this literature gap by conducting an economic evaluation of the heterologous effects of OPV, as one illustration of the potential efficiency of introducing LAVs.

OPV is used in many parts of the world as it is safe and effective at protecting children against lifelong polio paralysis. Past studies have demonstrated OPV’s and non-pathogenic enteroviruses’ strong heterologous effects against acute respiratory diseases induced influenza and other viruses, and OPV’s reduction in infant and child mortality.(7–9) Beyond the health benefits, OPV is attractive because it is safe (<1 adverse event per million vaccinees), inexpensive (<US$0.20 per dose), does not require trained medical staff (oral administration), and exists in three serotypes that could be used sequentially to extend the protection.(3)

We studied the benefit-cost and cost-effectiveness of OPV in two settings. The first setting is in a high child mortality setting, where multiple studies, in both natural experiments and randomized controlled trial settings, found that OPV reduced child mortality and morbidity by 10-36%.(8) The second setting is amidst the coronavirus-2019 (COVID-19) pandemic in a lower middle-income setting, where the availability of SARS-CoV-2-specific vaccines (hereinafter referred to as COVID-19 vaccines) is delayed due to development, manufacturing, and supply chain delays.(10) Even in the case of a pathogen for which the development of a vaccine is not particularly difficult, as was the case for SARS-CoV-2, there is still a wait of at least one year before a safe and effective vaccine can be deployed. The delay in immunization is further extended in lower-income countries: as of writing (early 2022), less than 10% of people in low-income countries have received at least one dose.(11) Even after administration, COVID-19 vaccines require several weeks until they become fully effective. The emergence of antigenically altered variants of SARS-CoV2 may further undermine the effectiveness of specific coronavirus vaccines.(12) In contrast, existing LAVs will likely face little delay as they have a proven track record of safety and can induce innate immunity almost immediately leading to protection against a broad range of viruses and variants.(9) Recently completed and ongoing studies explore how existing vaccines might help with the current pandemic, and as importantly, to prepare for future pandemics.(4,13,14) For OPV, to date, there are two ongoing trials investigating the effect of OPV against COVID-19.(15,16) The Russian study of 600 participants recently reported efficacy of OPV against COVID-19 incidence, while the preliminary results from Guinea-Bissau are not yet available. The analysis below uses plausible and conservative parameter values and sensitivity analysis to assess the economic attractiveness of these uses for OPV.

## Methods

### Setting 1: OPV against infant and child mortality in a high mortality setting

#### Model overview

We start with a birth cohort of one million in Guinea-Bissau, where the death rate in 2019 among children under five years of age was 78 per 1000 live births. In this setting, we assume all newborns receive the standard one dose of OPV at birth and three OPV doses at 6, 10, and 14 weeks of age together with other routine vaccination.(17) We model the impact of three cycles of an annual national immunization day campaign, in which eligible children (i.e., 100% coverage) receive one bi-valent OPV campaign dosage each year for three consecutive years, compared to a hypothetical cohort without this. Across all studies, the relative risk of all-cause mortality comparing after and before OPV campaigns among children is 0.75 (95% CI 0.69-0.82).(8) Conservatively, we headline one of the lowest estimates in the literature and set the relative risk of all-cause mortality following OPV at 0.90 [0.49-0.94] for the first dose for 0–11-month-old’s, and 0.92 [0.81-0.92] for each additional dose.(8,17,18) We estimated the overall intervention effectiveness and deaths averted per thousand doses, a metric commonly reported to assess vaccine effectiveness.(1) Analyses were conducted and reported for a range of estimates of vaccine effectiveness and in a range of values of child mortality where the intervention could be introduced.

#### Cost-effectiveness and benefit-cost analyses

All costs are reported in 2020 USD. Costs for the three OPV campaigns include vaccine, delivery, and wastage costs. Each child receives three doses of bivalent OPV, each costing $0.15 [0.12-0.19].(19) We searched for relevant campaign delivery costs, converted them to 2020 USD, and applied the average of $1.03 [0.72-1.56] per person per campaign and 10% [1-25] wastage rate (appendix p3).(20) The value per statistical-life (VSL) approach was taken to estimate the monetary value of mortality reduction, and standardized sensitivity analyses were undertaken in accordance with current guidelines (appendix p4).(21) Furthermore, since the intervention population is young children, we reduced the VSL by 50% to account for the fact that in some literature a lower VSL is suggested for young children.(22,23) We used the lowest VSL for the main result.

### Setting 2: OPV against COVID-19 in India

#### Model overview

A susceptible-exposed-infectious-recovered (SEIR) model was developed to estimate the population benefits of different vaccine scenarios. We adapted the model to more accurately reflect the transmission dynamics of COVID-19 (appendix pp 7-8). First, the “infectious” compartment was expanded into asymptomatic, symptomatic, hospitalized, and intensive care, to reflect different disease severity. We created two additional parallel sets of compartments for vaccinated individuals, reflecting the reduction in transmission and disease severity with vaccination. We assumed a fixed population size and did not incorporate background mortality or entry. We ran the model for 365 days.

#### Model calibration

The first wave of the COVID-19 epidemic among adults in India was selected to illustrate the potential use of OPV during a pandemic wave. India was chosen because it has national seroprevalence surveys and faced delays in immunizing a large proportion of the population with the COVID-19 vaccine. Following the national surveys, we set the first day of the simulation to mid-May 2020, when 0.7% of the population was estimated to be sero-positive and set 25% of them as the initial exposed population.(24–26) We calibrated the model so that similar levels of total infections, symptomatic cases, infection-fatality rate (IFR), and peak of the daily reported cases matched the observed (appendix p10-12). Since February 2021, India has entered a second wave that is far exceeding the severity of the first wave, while only 7% of the population has been vaccinated against COVID-19.(27) This analysis does not model the second wave because, in theory, it would have been prevented with both vaccination scenarios modeled described below, and therefore should not impact the estimated incremental benefits and costs between these scenarios.

#### Model scenarios

Two intervention scenarios with co-administration of the COVID-19 vaccine and OPV were studied (Table 1 and appendix p15). In both scenarios, the COVID-19 vaccine is introduced with a long implementation delay to reflect the time required to develop, test, approve, manufacture, procure, and administer. It also has a lag time between when the vaccine is administered and when it achieves full effect (about 3-5 weeks). In the first scenario, both vaccines are simultaneously administered on the same day, *t* days after the beginning of an epidemic wave. OPV is used to cover the 3-5 weeks before the COVID-19 vaccine achieves full effectiveness. In the second scenario, individuals expected to receive the COVID-19 vaccine will first receive OPV at day *t*, and then receive the COVID-19 vaccine *d* days after OPV administration (i.e., *t+d* days after the beginning of the wave). OPV is used as a bridge to fill this gap of *d* days. For both scenarios, the comparison scenarios include only the COVID-19 vaccine administered on the same day as the intervention scenario.

**Table 1.** Two immunization schedules adding OPV to COVID-19 vaccine.

| Schedule | Timing* |
| --- | --- |
| Simultaneous administration | COVID-19 vaccine and OPV both administered $t$ days after beginning of a wave |
| COVID-19 vaccine delayed | OPV administered $t$ days after beginning of wave and COVID-19 vaccine becomes available with a delay of $d$ days after OPV administration (i.e., $t+d$ days after beginning of wave) |

\* Calculations assuming administration of vaccines on days $t$ and $t+d$ , and that this approximates gradual coverage growth centered on those days.

#### Input parameters

Large-scale clinical studies of OPV showed that it was effective against influenza virus infection, with 1.9-5.8-fold reduction in morbidity.(28) Weighted average across these studies yields OPV effectiveness of 64%, with the lowest estimated at 47% (appendix p16). To be conservative, we assume OPV to be 20 [5-64]% immediate effectiveness against infection and severe illness (not against reducing infectivity among people who are infected) for COVID-19, slightly lower than the assumptions made by others.(15) For COVID-19 vaccines, we model the effect of a single-dose vaccine, similar to the vaccine produced by Johnson & Johnson, with 74 [65-95]% effectiveness against infections and reducing infectivity, and 95 [75-99]% effective against severity after 28 [7-35] days post-administration.(29) We further assume immunity from both vaccines last longer than the first wave. Vaccine coverage was set at 30 and 50% for both vaccines.

#### Outcomes of interest: Deaths averted per thousand immunized, cost-effectiveness and benefit-cost ratios per averted death

Deaths averted per thousand immunized (DATI) was estimated by dividing the number of deaths averted by the number of immunized adults. We assumed vaccine cost of $0.15 [0.12-0.19] and $10 [7.5-12.5] and delivery cost of $0.96 [0.56-1.56] and $1.49 [0.90-2.51] for one dose of OPV and COVID-19 vaccine, respectively (appendix p17). For both vaccines, we assumed a 10% [5-15] wastage rate. For benefit-cost analysis, we applied three sets of VSL estimates following the reference case standardized sensitivity analysis recommendation, extrapolating from the US VSL with Indian GNI per capita in 2019 at $6,920, and used the lowest VSL for the main result (appendix p24). 95% uncertainty ranges were calculated by incorporating reasonable ranges of a set of parameters (costs, COVID-19 vaccine effectiveness, epidemic) and repeating all calculations 1000 times using one draw of each parameter at each iteration (appendix p24).

#### Sensitivity analysis

For both settings, given the heterogeneity and uncertainty surrounding several input parameters, we ran a thorough set of sensitivity analyses for model inputs related to the epidemic (baseline mortality and severity parameters, basic reproduction number (R0), infectiousness ratio between symptomatic and asymptomatic individuals), vaccine (coverage, delay, effectiveness against infections, infectivity, severity), cost (vaccine, campaign, wastage), and VSL (GNI per capita) (appendix pp 5-6 and pp 24-33).

All analyses were conducted in R, version 4.0.1.

#### Role of the funding source

None

## Results

### Setting 1: OPV against infant and child mortality in a high mortality setting

Without the intervention, we would expect 50,000 deaths between ages 0-1, 6650 deaths between ages 1-2, and 6600 deaths between ages 2-3, reflecting the age-specific death rates in Guinea-Bissau (appendix p2). With the intervention, the number of total averted deaths with the campaigns is 5990 [4280-27590] deaths (6.0 [4.3-27.6] deaths averted per 1000 live births), which translates into a 9.5% [6.8-43.6] reduction in death from the scenario without the campaign. Deaths averted per 1000 campaign doses is estimated at 2.0 [1.4-9.2] (each child receives three doses). Total cost of the three-year campaign is estimated at US$3.9 [3.4-5.1] million. The cost-effectiveness ratio is approximately $650 [120-1240] per averted death, and the benefit-cost ratio is 110 [60-590] with a VSL of $71,000. Figure 1 depicts how these outcomes vary by background mortality and intervention effectiveness. With higher under-5 mortality rate and intervention effectiveness (upper-right quadrant), we would expect greater averted deaths per 1000 campaign doses (>2, Panel A), lower cost-effectiveness ratio (<$300, Panel B) and higher benefit-cost ratio (>200, Panel C). In contrast, this intervention may not be as cost-effective or cost-beneficial when the mortality rate or intervention effectiveness is lower (lower-left quadrant).

**Figure 1.**
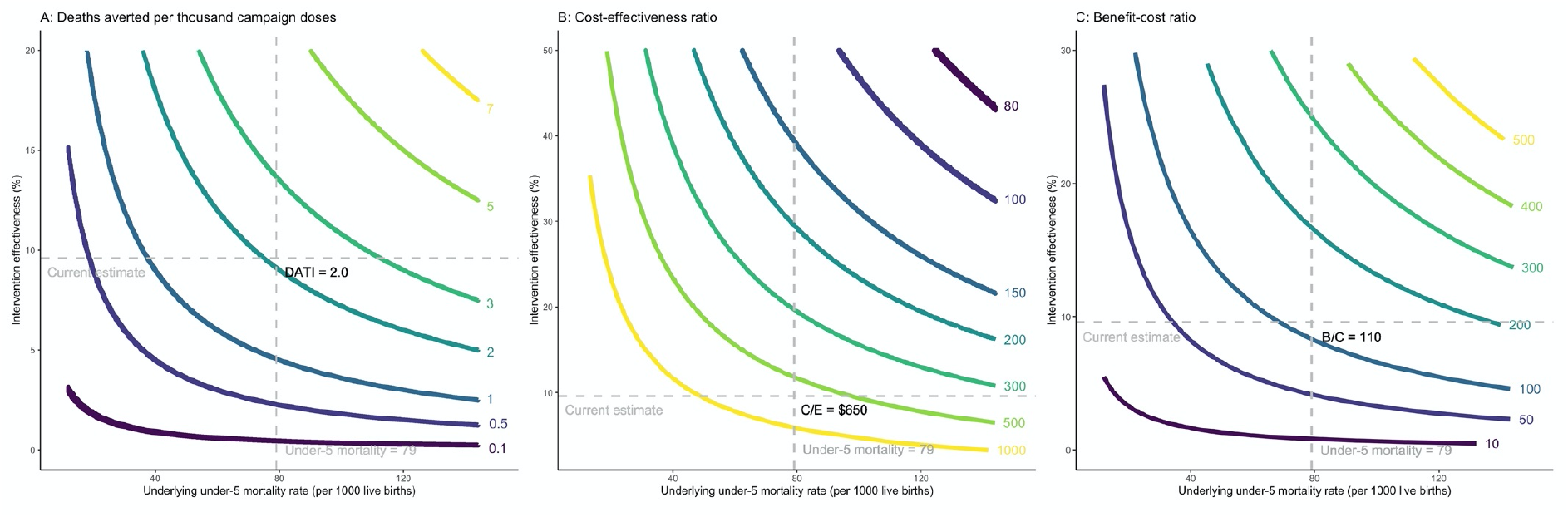
Cost-effectiveness and benefit-cost ratios per child death averted (by background mortality and intervention effectiveness) Headline assumptions about parameter values are denoted by the dashed lines. A: Incremental deaths averted per thousand campaign doses B: Incremental cost-effectiveness ratio per averted death C: Incremental benefit-cost ratio The intervention was studied among children under 3 years of age, consistent with the scientific literature, and the results are estimated based on under-3 mortality rates. Since under-5 mortality rate is a more commonly used indicator, we extrapolated the under-3 mortality rate to under-5 mortality rate in the x-axes by multiplying it by the ratio of global average of under-5 to under-3 mortality. Note on Panel A: Each curve shows combinations of the underlying under-5 mortality rate and intervention effectiveness that makes deaths averted per 1000 campaign doses (DATI) constant along the curve. For example, the yellow curve on the top right shows combinations that result in 7 deaths averted per one thousand campaign doses administered. Note on Panel B: Cost-effectiveness ratio (C/E) is expressed as cost per child death averted. For example, the yellow curve on the bottom left shows combinations that result in $1000 per child death averted. Note on Panel C: The benefit-to-cost ratio (B/C) is the ratio of the dollar value of benefits to the dollar value of costs. For example, the yellow curve on the top right shows combinations that result in a benefit-to-cost ratio of 500.

### Setting 2: OPV against COVID-19 in India

Without any vaccines, the model projects 25% of the population in this wave to have ever been infected over the course of 365 days (Table 1). At a vaccine coverage rate of 30%, simultaneously co-administering OPV with COVID-19 vaccine would yield approximately 13-35% reduction in infections and deaths if administered before day 100, and 2-13% if administered between day 100-200, compared to the scenario with only COVID-19 vaccine (appendix pp18-19).

The main results are presented in Table 2 and Figures 2 and 3. The figures reflect the outcomes under plausible but uncertain ranges for three key parameters – length of COVID-19 vaccine and OPV implementation delay and OPV effectiveness against COVID-19. When both vaccines are co-administered simultaneously, the most substantial incremental impact of OPV was estimated to occur around day 100 into the wave, corresponding to a few days before the peak of the wave. Assuming OPV effectiveness against COVID-19 of 20%, DATI was estimated at 0.05, and incremental cost-effectiveness and benefit-cost ratios per averted death at $23000 and 17 (with VSL $388,000), respectively. Even if both vaccines are administered at day 200 since the beginning of the wave, cost-effectiveness and benefit-cost ratios were estimated favorably at $65,000 per death averted and 6:1, respectively.

**Table 2.**
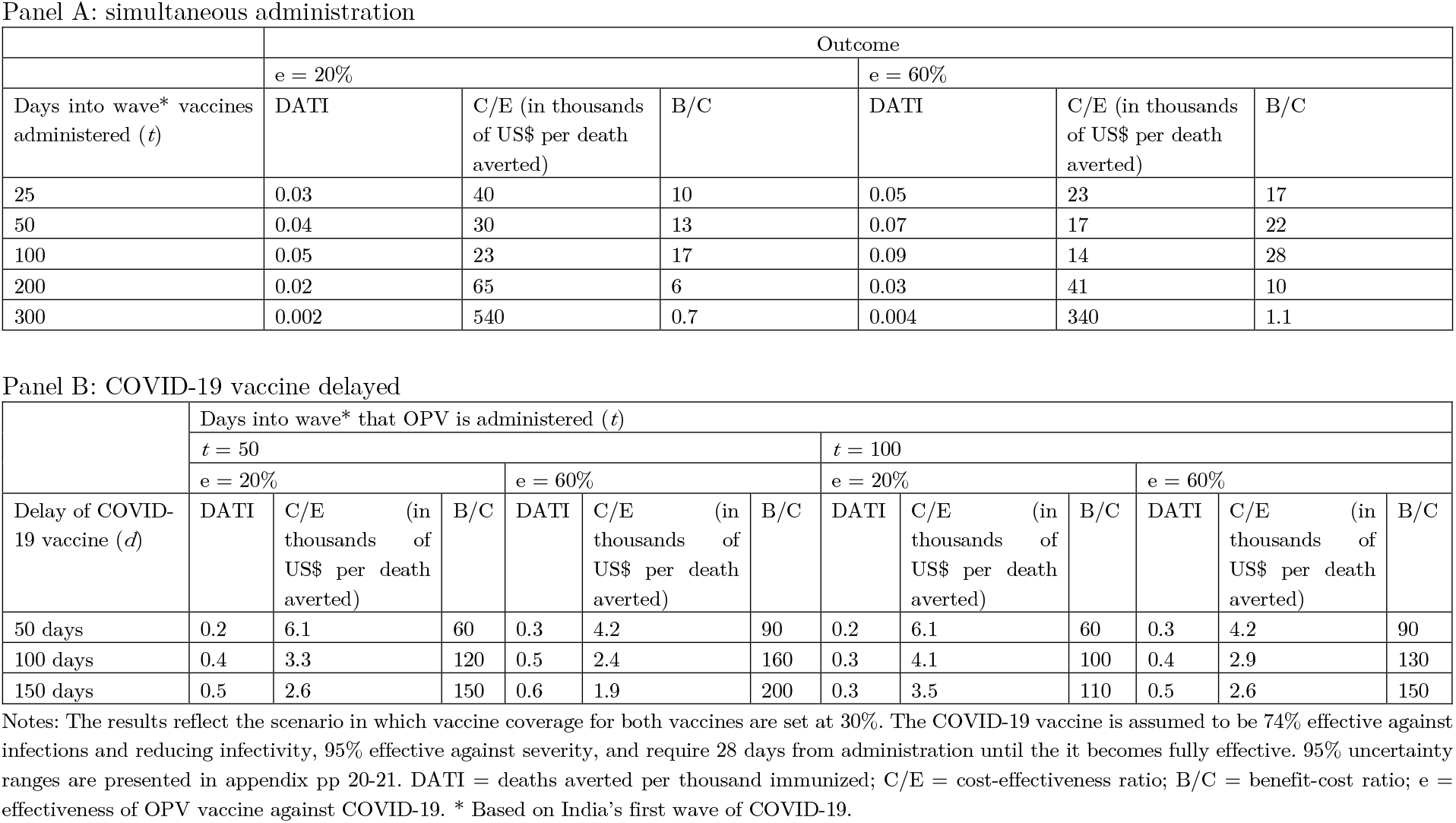
Outcomes of COVID-19 vaccine + OPV schedule.

**Figure 2.**
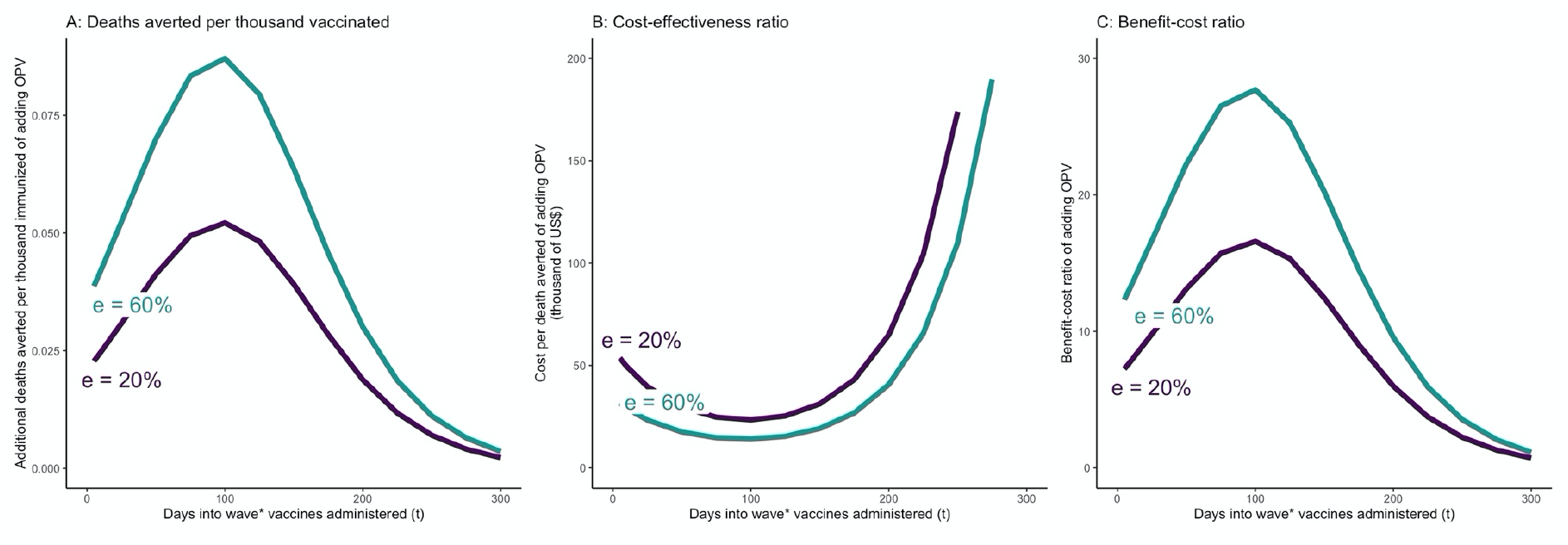
OPV and COVID-19 vaccine simultaneously administered *t* days after beginning of epidemic wave – incremental mortality impact, cost-effectiveness, and benefit-cost of adding OPV. A: Incremental deaths averted per 1000 individuals of adding OPV to COVID-19 vaccine only schedule B: Incremental cost-effectiveness ratio per averted death of adding OPV to COVID-19 vaccine only schedule C: Incremental benefit-cost ratio of adding OPV to COVID-19 vaccine only schedule The results reflect the scenario in which vaccine coverage for both vaccines are set at 30%. OPV is administered simultaneously with the COVID-19 vaccine on day *t*. The COVID-19 vaccine is assumed to be 74% effective against infections and reducing infectivity, 95% effective against severity, and require 28 days from administration until the it becomes fully effective. e = effectiveness of OPV vaccine against COVID-19.

**Figure 3.**
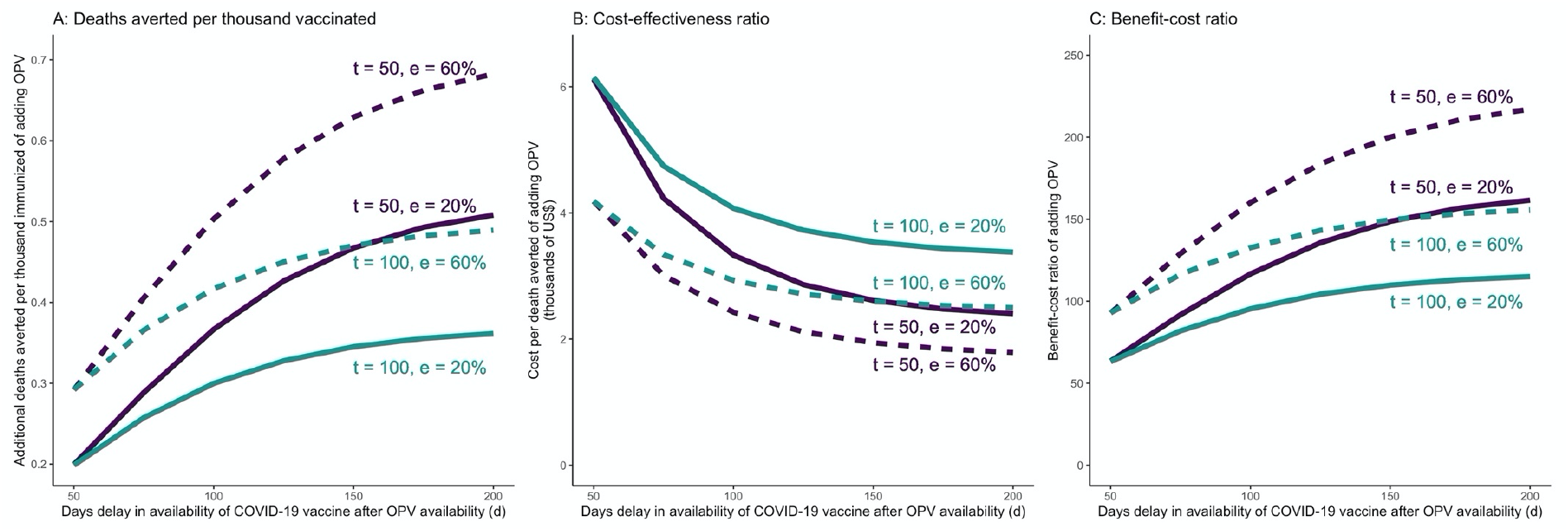
COVID-19 vaccine is delayed in a COVID-19 vaccine + OPV schedule – incremental mortality impact, cost-effectiveness, and benefit-cost of adding OPV as a function of delay in COVID-19 vaccine availability. A: Incremental deaths averted per 1000 individuals of adding OPV to COVID-19 vaccine only schedule B: Incremental cost-effectiveness ratio per averted death of adding OPV to COVID-19 vaccine only schedule C: Incremental benefit-cost ratio of adding OPV to COVID-19 vaccine only schedule The results reflect the scenario in which vaccine coverage for both vaccines are set at 30%. OPV is administered on day *t*, and the COVID-19 vaccine is administered *d* days after OPV. The COVID-19 vaccine is assumed to be 74% effective against infections and reducing infectivity, 95% effective against severity, and require 28 days from administration until the it becomes fully effective. e = effectiveness of OPV vaccine against COVID-19.

When the COVID-19 vaccine is delayed, larger time gaps between administering OPV and COVID-19 vaccine would lead to more favorable incremental outcomes for OPV. Assuming OPV is introduced 50 days after the beginning of the wave, even if OPV has low effectiveness against COVID-19 (at 20%), we would expect DATI of 0.2-0.5, incremental cost-effectiveness and benefit-cost per averted death of $2600-6100 and 60-150, respectively (Table 2). A later introduction of OPV (100 days since the beginning of the wave) would yield less preferable but still acceptable outcomes: DATI of 0.2-0.3, incremental cost-effectiveness of $3500-6100 per death averted and a benefit-to-cost ratio in the range of 60-110. Results for higher OPV effectiveness (60%) and vaccine coverage (50%) are presented in Table 2 and appendix pp 20-21.

#### Sensitivity analysis

The full results from the sensitivity analyses are presented in the appendix. In setting 1, the model is most sensitive to plausible variations in baseline mortality, OPV effectiveness, campaign delivery cost, and GNI per capita (appendix p6; Figure 2 shows the results for plausible ranges of mortality and effectiveness). In setting 2, the model is most sensitive to variations in some of the underlying epidemic parameters, vaccines’ effectiveness delay, and OPV delivery cost (appendix pp 24-33; figure 2 and 3 show the results for plausible ranges of implementation delay and OPV effectiveness).

## Discussion

This study complements existing and ongoing studies that have demonstrated the high effectiveness of LAVs in reducing disease burden through activation of innate immune responses. Our economic evaluation points to the potential attractiveness of LAVs, such as OPV, as highly cost-effective and cost-beneficial interventions against child mortality. In the context of COVID-19, the results support the call for clinical investigations to explore its potential against both COVID-19 and future pandemics, especially the co-administration strategies alongside the COVID-19 vaccine.

Nonetheless the study has a number of limitations First, while the effectiveness of OPV in preventing a variety of respiratory infections has been documented, to date, there is no empirical estimate on the effect of OPV in reducing transmission of SARS-CoV-2 or severity of COVID-19. A recent randomized trial of BCG vaccination, another LAV, found that BCG revaccination decreased COVID-19 risk by 68% among the older adults, whereas a press release from another randomized trial testing a primary dose of BCG among older adults reported no effect on COVID-19 incidence.(14,30) Biologically and epidemiologically plausible evidence suggest that there may be a beneficial effect of OPV in relation to COVID-19.(9,31) We therefore applied a wide range of effectiveness estimates in our sensitivity analysis to account for this uncertainty. If future clinical trials show a zero effect of OPV against COVID-19, then the analyses above would be superfluous.

Second, our SEIR model is basic and does not account for more nuanced aspects of the dynamic. For example, it does not account for heterogeneity in population mixing patterns nor model more than one epidemic wave. However, if the vaccines are deployed during the first wave, as we have assumed, the following waves should not impact our results because of our focus on the marginal benefits and costs between scenarios. Other model limitations include our assumptions that the duration of immunity for both vaccines last longer than the length of one epidemic wave (which may or may not be accurate), and a flat vaccine coverage rate across the population. The model was calibrated against data from India’s first wave as an illustration and we by no means aimed to accurately project the actual epidemic. India’s first wave was also relatively mild in comparison to other countries (appendix p13), so the estimated health impacts are likely conservative. Similarly, income levels, vaccine and delivery costs will lead to differences in benefit-cost and cost-effectiveness ratios. In the child mortality analysis, we applied the mortality rates and effect sizes from studies conducted in Guinea-Bissau, and these assumptions may not be applicable in other settings, such as environments with much lower mortality, higher income and costs, or with different vaccine schedules.

Third, our study does not address an important point of discussion relating to the debate on global OPV cessation, set to be executed in 2024.(32) On rare occasions OPV causes vaccine-associated paralytic polio (VAPP), most commonly among immunocompromised individuals. Although the risk is real, its probability is vanishingly small – less than one in a million. This point and explicit comparisons with inactivated polio vaccines need to be brought into further analyses. We did not explicitly incorporate potential negative health consequences (such as VAPP) in our analysis. Qualitatively these considerations are unlikely to impact our main results. In particular, our findings suggest that, in the case of child mortality, the trade-off would be between averting 6,000 child deaths versus avoiding one possible case of VAPP. Policymakers should be aware of these trade-offs, and we believe this study serves as an important contribution to this ongoing debate. Finally, more analysis is needed on supply constraints of OPV and how to resolve them.(33)

Fourth, beyond the use of LAVs modeled in this study, there are further modes of use of LAVs to contribute to the reduction of child mortality and pandemic burden. These are summarized in Table 3. More comprehensive analyses will include one or more of these additional modes of use.

**Table 3.** Potential uses of live attenuated vaccines in the context of childhood immunization and COVID-19.

| Setting 1: childhood immunization | Setting 2: COVID-19 |
| --- | --- |
| 1. Modification of some non-live vaccines (such as DTP, Pentavalent) <sup>38</sup> | 1. “Bridge” use until COVID vaccine becomes available for the general population, responders (medical, fire, police), and at-risk populations (hourly workers, gig workers, undocumented residents, etc) |
| 2. Enhancement of maternal immune system with administration in adolescence or fertile age to secure better heterologous responses among their offspring when vaccinated with the same vaccine<sup>39</sup> | 2. As an adjuvant concomitant to administration of a COVID vaccine to provide incremental benefit; to boost responses in elderly or individuals with comorbidity |
| 3. Further reduction of mortality risk with additional dose of LAVs (either as booster or campaign), or with combinations of LAVs<sup>40</sup> | 3. Ring use as a quick response to appearance of infection in pre- or post-pandemic flare-ups in the general population, institutionalized elderly and caretakers, prisoners and guards, and other institutionalized groups |
| 4. Use of LAVs in high-risk groups like premature and low birth weight infants<sup>41</sup> | 4. Therapeutic use (hypothetical) for infected individuals soon after detection of infection to reduce disease severity<sup>42</sup> |
| 5. Instead of switching from OPV to IPV in childhood immunization, schedule switch to IPV/OPV mix (like China today) |  |

In the child mortality setting, we found that a 9.5% campaign effectiveness against child mortality results in cost-effectiveness and benefit-cost ratios of $650 and 170 per averted death. To put these numbers in perspective, previous studies found the cost-effectiveness ratio of national immunization programs to be in the order of $300-1800 per averted death.(34) GAVI, the Vaccine Alliance, estimated the benefit-cost ratio for ten antigens to be around 50-70 by 2030.(2) Even with lower OPV campaign effectiveness, for example, 5-10% against child mortality, the cost-effectiveness and benefit-cost ratios per averted death are economically attractive relative to other interventions (Figure 1). Deaths averted per thousand vaccinated was estimated at 2 per campaign dose (or 6 per vaccinated child), which is comparable to a recent study that estimated deaths averted per thousand vaccinated across ten antigens to be 3.5 in 98 countries.(1) It is important to note that mortality reduction is additive to, not a substitute for, mortality reduction in the standard immunization schedule.

In the COVID-19 setting, co-administering OPV alongside the COVID-19 vaccine yielded favorable economic results. Applying OPV for bridge use becomes more attractive with longer COVID-19 vaccine delays, shorter OPV delay, and higher OPV effectiveness against the disease (Figures 2,3). In line with other studies, we found that the cumulative infections and deaths are highly sensitive to the time delay of the COVID-19 vaccine rollout.(35) In other words, the timing of administering vaccines with some level of effectiveness against COVID-19 is in the short term (and especially at the beginning of the epidemic) more important than its effectiveness. Further considering large macroeconomic and social consequences related to pandemic control measures could yield even larger benefits for introducing bridge interventions measures.(36) In comparison, one US-based study estimated that COVID-19 vaccine has an average incremental cost of $8200 per QALY gained for adults (corresponding to a cost of several hundred thousand dollars per averted death).(37) Empirically testing whether readily available LAVs could bridge the gap until COVID-19 vaccines are available through clinical trials could potentially lead to mitigating further damage from consequent waves and future pandemics.

## Conclusion

Economic evaluation points to the large potential of LAVs, such as OPV, to add a significant new dimension to the ongoing efforts to reduce child mortality. LAV action against COVID-19 remains to be shown in trials, but this economic evaluation shows sufficiently attractive potential to place high priority on the relevant trials.

## Data Availability

All data produced in the present study are available upon reasonable request to the authors

## Acknowledgement

We would like to acknowledge Drs. Robert Gallo, Walter Orenstein, Kelly Sanders, Jaime Sepulveda, Kimberly Thompson, and Damian Walker for their scientific inputs. Conclusions and views expressed in the paper are those of the authors alone.

## Authors’ contributions

AYC, SMB, and DTJ conceptualised the study and contributed to model development. AYC collected the data and ran the analyses. AYC, SMB, and DTJ interpreted the results. AYC and DTJ wrote the manuscript. All authors reviewed and edited the manuscript. The corresponding author had final responsibility for the decision to submit for publication.

## Conflict of interest

Dr. Netea reports activities outside the submitted work from Trained Therapeutix Discovery; in addition, Dr. Netea has a patent Inhibitors of trained immunity licensed, and a patent Stimulators of trained immunity licensed. Dr. Kottilil reports support outside the submitted work from Regeneron Pharmaceuticals, grants from Gilead Sciences, grants from Merck Inc, grants from Arbutus Pharmaceuticals, during the conduct of the study.

## Author approval

all authors have seen and approved the manuscript.

## Role of funding source

No funding

## Ethics committee approval

Not applicable

## Notes

**Funding:** The contribution by DTJ was supported through grants from Trond Mohn Foundation (BFS2019MT02) and Norad (RAF-18/0009) through the Bergen Centre for Ethics and Priority Setting.

### Funding Statement

This study did not receive any funding

